# C-reactive protein as a triage tool for adults with presumptive pulmonary tuberculosis in South Africa: a prospective cohort study

**DOI:** 10.1101/2021.10.12.21264791

**Authors:** Claire J Calderwood, Byron WP Reeve, Tiffeney Mann, Zaida Palmer, Georgina Nyawo, Hridesh Mishra, Ibrahim Abubakar, Mahdad Noursadeghi, Grant Theron, Rishi K Gupta

## Abstract

**Background:** Identification of an accurate, low-cost triage test for pulmonary TB among people presenting to healthcare facilities is an urgent global research priority. We assessed the diagnostic accuracy and clinical utility of C-reactive protein (CRP) for TB triage among symptomatic adult outpatients, irrespective of HIV status.

**Methods:** We prospectively enrolled adults reporting at least one (for people with HIV) or two (for people without HIV) symptoms of cough, fever, night sweats, or weight loss at two TB clinics in Cape Town, South Africa. Participants provided sputum for culture and Xpert MTB/RIF Ultra. We evaluated the diagnostic accuracy of CRP (measured using a laboratory-based assay) against a TB-culture reference standard as the area under the receiver operating characteristic curve (AUROC) and sensitivity and specificity at pre-specified thresholds. We assessed clinical utility using decision curve analysis, benchmarked against WHO recommendations.

**Results:** Of 932 included individuals, 255 (27%) had culture-confirmed TB and 389 (42%) were living with HIV. CRP demonstrated an AUROC of 0.80 (95% confidence interval 0.77–0.83), with sensitivity 93% (89–95%) and specificity 54% (50–58%) using a primary cut-off of ≥10mg/L. Performance was similar among people with HIV to those without. In decision curve analysis, CRP-based triage offered greater clinical utility than confirmatory testing for all up to a number willing to test threshold of 20 confirmatory tests per true positive TB case diagnosed.

**Conclusions:** CRP approached the WHO-defined minimum performance for a TB triage test and showed evidence of clinical utility among symptomatic outpatients, irrespective of HIV status.

## Background

Of the estimated 10 million people who developed tuberculosis (TB) in 2019, 2.9 million were not reported to TB programmes. Delayed or missed diagnoses result in worse outcomes for individuals and ongoing community transmission.^1^ The World Health Organization (WHO) has emphasised the urgent need for better tools to identify people with TB, in order to find these ‘missing millions’. A key priority is development of a rapid, point-of-care, low-cost, non-sputum-based triage test with high sensitivity to target more expensive or resource-intensive confirmatory testing. WHO proposed that such a triage test should be deployable even at lowest-level health facilities and achieve at least 90% sensitivity and 70% specificity or, optimally, 95% sensitivity and 80% specificity.^2,3^

C-reactive protein (CRP) is an acute phase reactant commonly used in clinical practice as a non-specific marker of inflammation. It may be measured on finger-prick blood samples using point-of-care platforms at a cost of US$2 per test, with results available in minutes. These attributes meet many of the operational characteristics of a triage test recommended by WHO.^2^ Recent WHO guidance has recommended that CRP (with a threshold of >5mg/L) may be used to screen for TB among adults and adolescents living with HIV, following a meta-analysis which demonstrated sensitivity 90% (95% confidence interval [95%CI] 78– 96%) and specificity 50% (29–71%) among outpatients initiating anti-retroviral therapy.^4^ However, the value of CRP as a triage test among symptomatic people presenting to healthcare facilities, particularly in unselected populations with and without HIV, is less clear. In two systematic reviews, no identified studies prospectively evaluated CRP-based triage in symptomatic outpatients, without pre-selection based on HIV or smear status.^5–9^ One subsequent study has evaluated the diagnostic accuracy of CRP for pulmonary TB in a case-control design. The best performance was achieved using a threshold ≥12mg/L (sensitivity 85% [95%CI 80–88%] and specificity 70% [65–74%]), but clinical utility to guide confirmatory testing in real-world cohorts remains unclear.^10^

We evaluated the diagnostic accuracy and clinical utility of CRP for triage among symptomatic adults in a large prospective observational cohort recruited from two primary health clinic-based TB services in Cape Town, South Africa. Our objectives were to evaluate whether CRP meets the WHO-recommended diagnostic accuracy targets for a triage test and to assess the clinical utility of CRP to prioritize use of confirmatory testing for TB in high-TB burden settings, irrespective of HIV status.

## Methods

### Study population

Adults (≥18 years) presenting to two government primary health clinics and reporting at least one (for people with HIV) or two (for people without HIV) WHO-defined TB symptom of fever, cough (any duration if HIV-positive; >2 weeks if HIV-negative), weight loss or sweats in Cape Town, South Africa were consecutively invited to participate (April 2016 - November 2020). Participants provided informed written consent before undergoing evaluation as reported previously.^11,12^

### Laboratory procedures

Participants provided two sputum specimens for reference standard testing. The first underwent two Ziehl-Neelson (ZN) stains and microscopy, and a liquid TB culture (MGIT960); the second was tested with Xpert MTB/RIF Ultra (hereafter ‘Ultra’). Ultra was performed retrospectively on samples collected prior to its implementation [2016-2017] and as part of routine care from 2018 onwards.^12^ Sputum induction was conducted where required, as previously described.^11^ Participants who did not self-disclose that they were living with HIV at study enrolment were offered HIV testing; CD4 counts, and haemoglobin were extracted from clinic records where available. We classified anaemia per WHO definitions (≤12g/dL women, <13g/dL men).^13^ Sera were collected at the same visit and biobanked. CRP was measured in sera using the Cobas high-sensitive immunoturbidimetric assay (Roche Diagnostics Limited, Burgess Hill, UK), masked to clinical details and reference standard test results.

### Outcome definition

For our primary analysis, participants with TB were defined as those with positive liquid culture for *Mycobacterium tuberculosis* complex (confirmed by GenoType MTBDR*plus* [Hain Lifescience GmbH, Nehren, Germany]). Participants with missing CRP or missing or indeterminate culture results were excluded from the analysis.

### Diagnostic accuracy analysis

This study is reported in accordance with the Standards for Reporting Diagnostic Accuracy Studies guidelines. All analyses were conducted in the statistical computing environment R (version 4.0.2). Our sample size calculation is shown in **Supplementary Figure 1**.

We investigated the discriminatory ability of CRP overall and stratified by HIV status and history of previous TB, reflecting important risk factors for TB in our population which we hypothesised may modify the diagnostic accuracy of CRP. In further pre-specified analyses, we stratified by age, sex, smoking status, CD4 count, anti-retroviral therapy status (ART), time since previous TB, and indices of disease severity at presentation (presence of anaemia, underweight [classified by BMI], and TB Score II severity category^14^).

Receiver operating characteristic (ROC) curves were constructed using the ‘pROC’ package.^15^ Confidence intervals of the area under the ROC curve (AUROC) were calculated using the DeLong method.^16^ For each subgroup analysis, stratum-specific AUROC were calculated and compared using DeLong tests. We also examined correlations between CRP and measures of disease severity among participants with TB (haemoglobin, body mass index, TB Score II, and culture days to positivity) using scatterplots and Spearman rank correlation.

Sensitivities, specificities and predictive values of CRP overall, and stratified by key subgroups of interest, were calculated at a primary threshold of 10mg/L, as proposed previously.^17,18^ We assessed a range of alternative cut-offs as secondary analyses, including: (a) CRP >5mg/L, consistent with recent WHO guidance for TB screening among PLHIV^4^; (b) the maximum Youden index; and (c) fixing sensitivity at 95% (reflecting the optimum sensitivity recommended for a triage test by WHO).^4^

We have previously reported false positive Ultra results in our high-burden setting.^11^ We therefore evaluated the potential of CRP to discriminate the culture status of Ultra-positive participants, including those with ‘trace’ results. We also evaluated the diagnostic accuracy of CRP for Ultra-negative or smear-negative, culture-positive TB to explore the potential for CRP to detect Ultra- or smear-negative cases. For a subset of the study population with data available, we stratified analyses by whether participants were able to spontaneously expectorate sputum, to assess the potential role for CRP to direct use of sputum induction.

### Clinical utility analysis

To further assess the potential clinical utility of CRP as a triage test, we calculated positive and negative predictive values for TB and performed decision curve analysis using the ‘rmda’ package in R.^19,20^ Decision curve analysis quantifies ‘net benefit’ as the proportion of true positives minus false positives, weighted by the ‘threshold probability’ for confirmatory testing. The threshold probability reflects the cost: benefit ratio of confirmatory testing and can also be expressed as a ‘number willing to test’ with a confirmatory assay to detect one true positive TB case (the inverse of the threshold probability). In decision curve analysis, the optimal approach is the one with highest net benefit across a clinically relevant number willing to test range. For TB the number willing to treat range will be context-specific, influenced by individual and community-level costs associated with TB, and resources available for TB diagnosis. It is likely to vary across health services and world regions.

We examined the net benefit of using CRP (with a cut-off of ≥10mg/L) to guide confirmatory testing across a number willing to test range from five to 100 (corresponding to threshold probabilities of one to 20%) and compared this to alternative strategies of offering confirmatory testing to all or confirmatory testing to none. In our study population, ‘confirmatory testing for all’ is equivalent to testing all individuals presenting to a clinic who meet WHO criteria for TB evaluation with a TB culture.

We also compared CRP performance to hypothetical dichotomous biomarkers simulated to meet WHO-recommended optimum (sensitivity 95%; specificity 80%) and minimum (sensitivity 90%; specificity 70%) diagnostic accuracy criteria for a triage test. We simulated these performance metrics by randomly sampling the relevant proportion of true positive and false positive results in the study population and allocating positive results to these sampled individuals, with the remainder allocated as negative.

### Sensitivity analyses

We performed a range of sensitivity analyses. First, we explored alternative reference standard definitions, using (a) culture- or Ultra-positive (including Ultra-trace) TB; and (b) Ultra alone. We repeated the decision curve analysis using an Ultra reference standard, given that Ultra is frequently the only confirmatory test available in routine care.

We also explored the potential impact of population TB prevalence on our findings by simulating different disease prevalences (range 5-40%) for predictive value estimates and decision curve analyses (using the rmda package^19,20^). In the latter analysis, we explored the number willing to test range in which applying CRP appeared to be the strategy with highest net benefit, compared to confirmatory testing for all or none, to assess its potential utility for triage across a range of settings.

### Ethics

This study was approved by the Stellenbosch University Faculty of Health Sciences Research Ethics Committee (N14/10/136).

## Results

### Study population

Between April 2016 and November 2020, 1,097 participants were recruited; 932 (85%) had CRP and culture results available and were included in the primary analysis (**Figure 1**). Characteristics of excluded participants are in **Supplementary Table 1**; there were no systematic differences *versus* those included. Among participants, the median age was 36 years (IQR 28–47 years). 46% were female (n=430) and 42% were PLHIV (n=389), of whom 67% were receiving ART at enrolment (n=259; **Table 1**). Twenty-seven percent of the study population were TB culture-positive (n=255), among whom the median time to positive culture was eight days (IQR 6–12 days, n=249 with data available).

**Figure 1:**
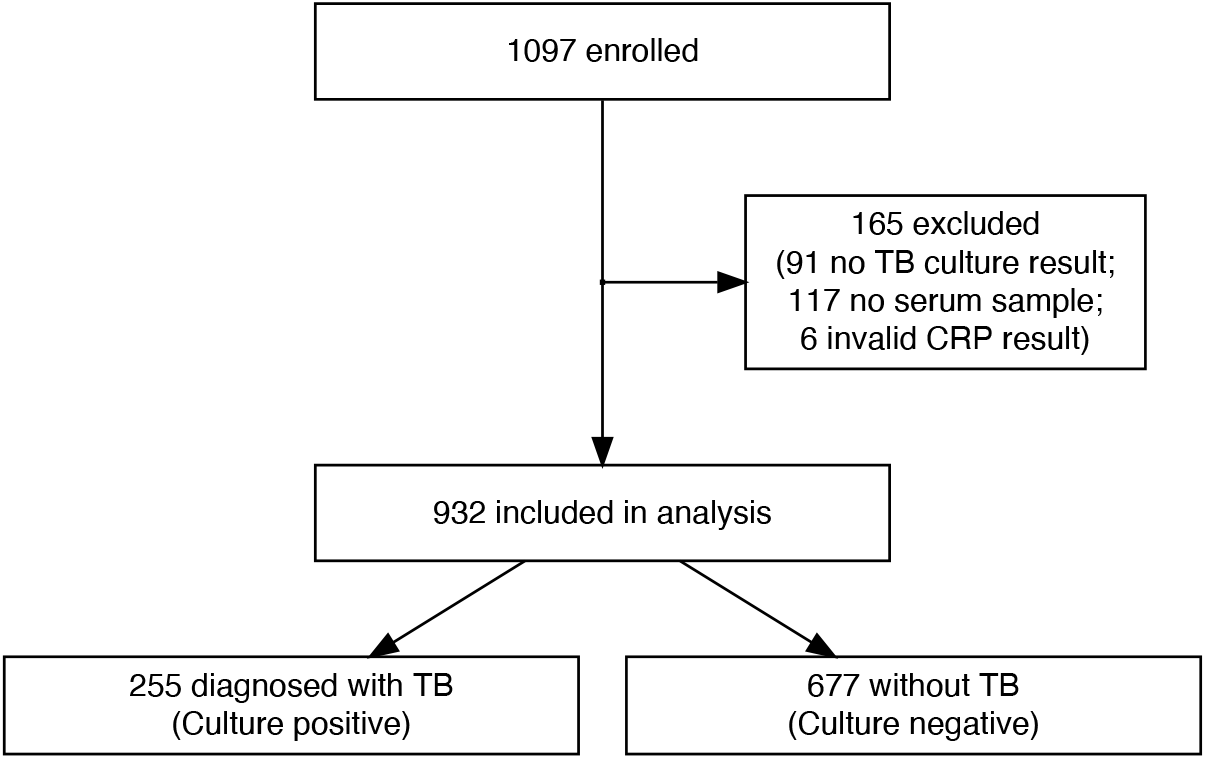
Overview of study cohort.

**Table 1:**
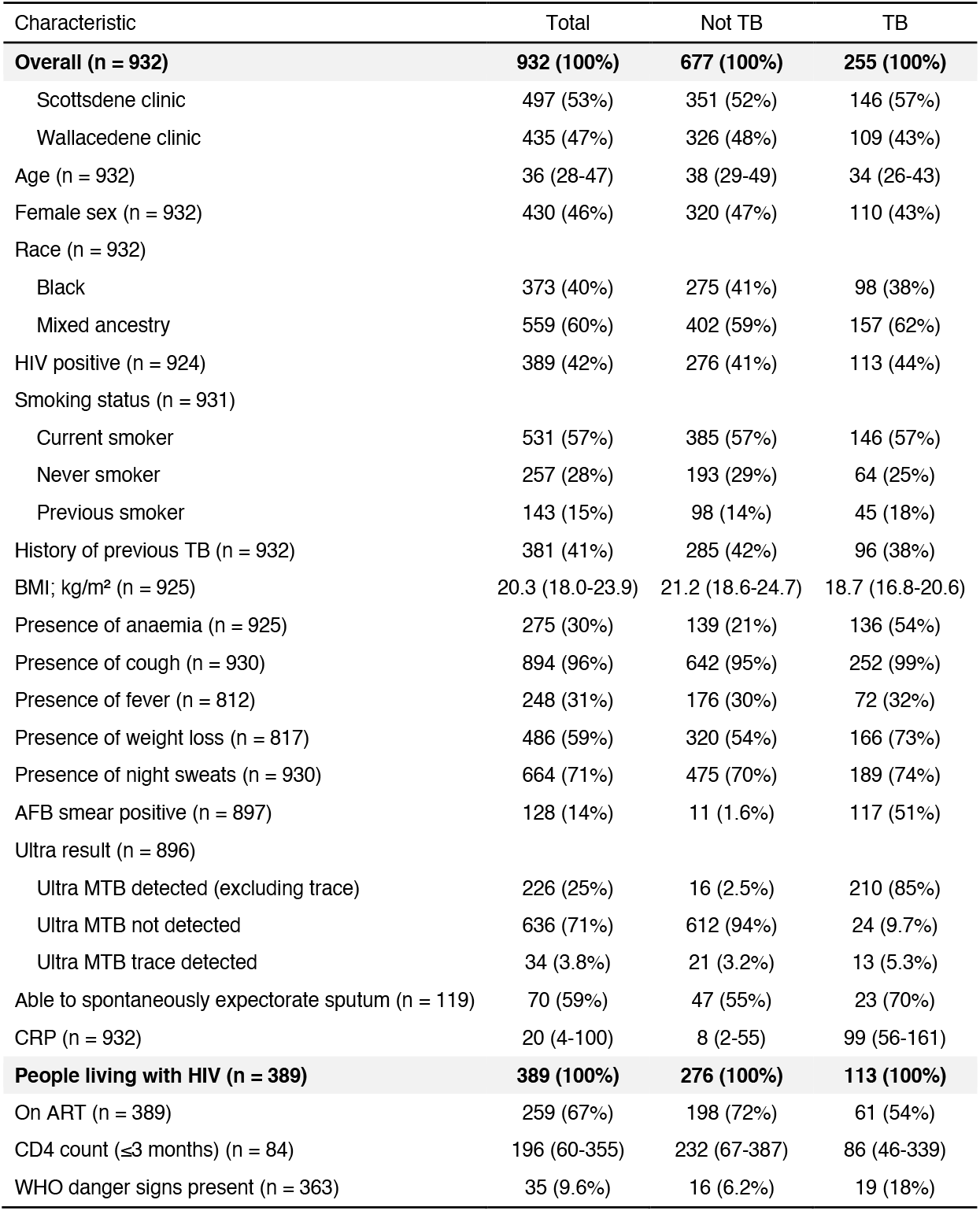
Characteristics of study population. Data are n (%) for categorical variables or median (interquartile range) for continuous data. Anaemia was defined as per WHO: haemoglobin ≤12g/dL for women and <13g/dL for men. **Abbreviations:** ART: antiretroviral therapy, BMI: body mass index, CRP: C-reactive protein, WHO: World Health Organization.

### Diagnostic accuracy

The overall AUROC of CRP for culture-positive TB was 0.80 (95%CI 0.77–0.83). This did not differ by HIV or previous TB status (DeLong test p=0.1 and p=0.5 respectively; **Figure 2**). AUROCs did not appear to be affected by age, ethnicity, smoking status, BMI, or time since last TB episode among those with previous TB (p≥0.1 across strata of each subgroup; **Supplementary Figure 2**). However, AUROCs were lower amongst people with anaemia compared to those without (0.68 [95%CI 0.62–0.75] vs. 0.80 [0.76–0.84]; p=0.003). There was some evidence of lower AUROC among men as compared to women (0.77 [0.73–0.81] and 0.84 [0.80–0.88] respectively; p=0.03).

**Figure 2:**
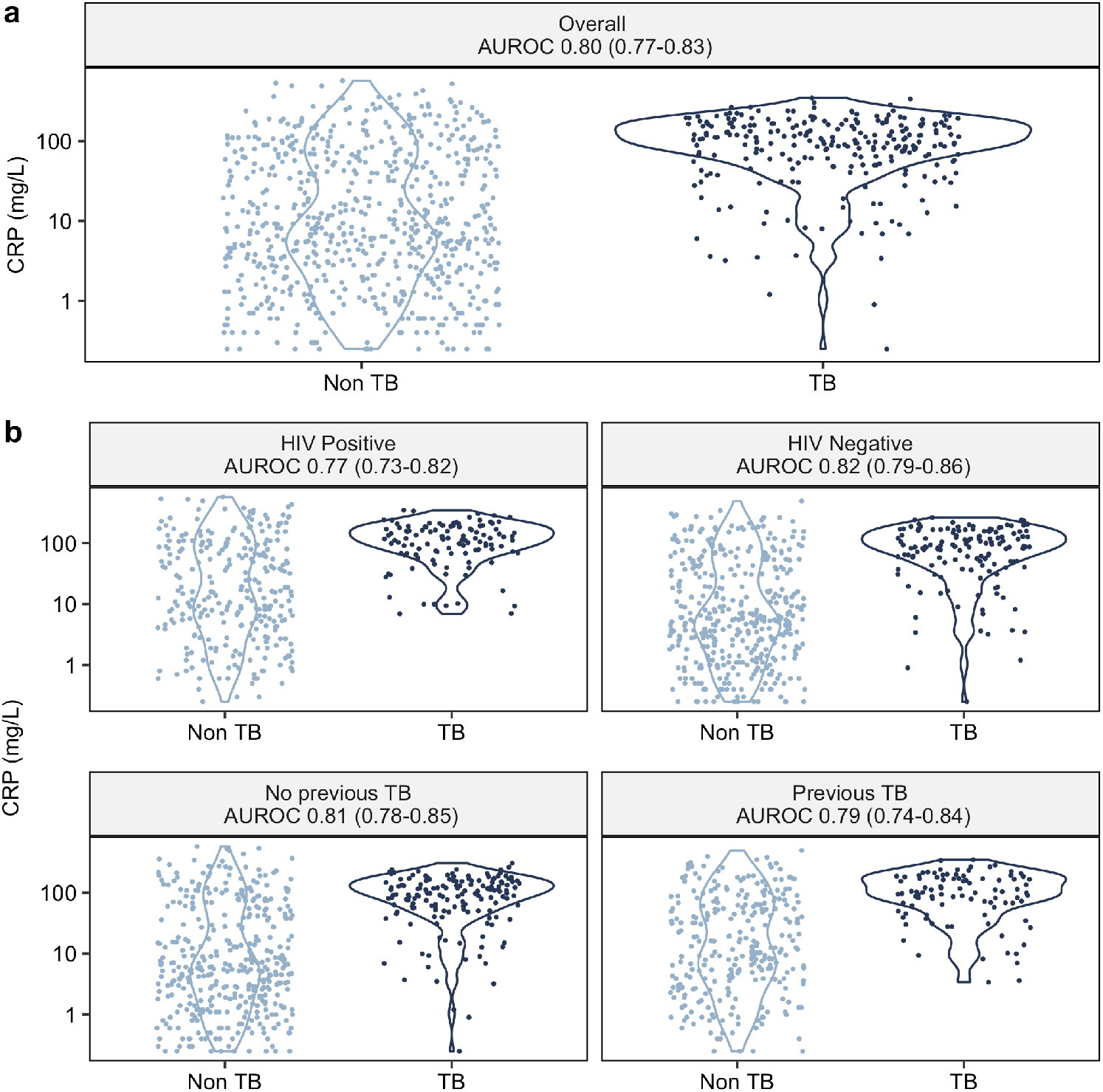
Discriminatory ability of CRP for culture-confirmed pulmonary TB among adults presenting to outpatient care with TB-related symptoms. Area under the receiver operator characteristic curve (AUROC) of CRP to distinguish pulmonary TB versus culture-negative in (a) whole study population and (b) key subgroups of interest; presented with CRP on a logarithmic scale. There was no difference in AUROC between HIV positive and HIV negative participants (DeLong test, p = 0.1) or among people with a history of previous TB compared to those without (p = 0.5).

Using our primary CRP threshold of ≥10mg/L, 546/932 (59%) of participants had a positive triage test. Sensitivity was 93% (95% CI 89–95%) and specificity was 54% (50–58%), thereby meeting the WHO-recommended minimum 90% sensitivity for a triage test, but not the minimum 70% specificity (**Table 2**). The numbers needed to test with culture to detect a true TB case were 2.3 and 20 among CRP-positive and CRP-negative participants, respectively. Using the >5mg/L cut-off, sensitivity was higher (97% [94–98%]), but specificity was only 39% (35–42%). At the maximum Youden index (CRP threshold ≥28mg/L), CRP approached both minimum WHO TPP criteria, with sensitivity 87% (95%CI 83–91%) and specificity 66% (62–69%; **Supplementary Table 2**).

**Table 2:**
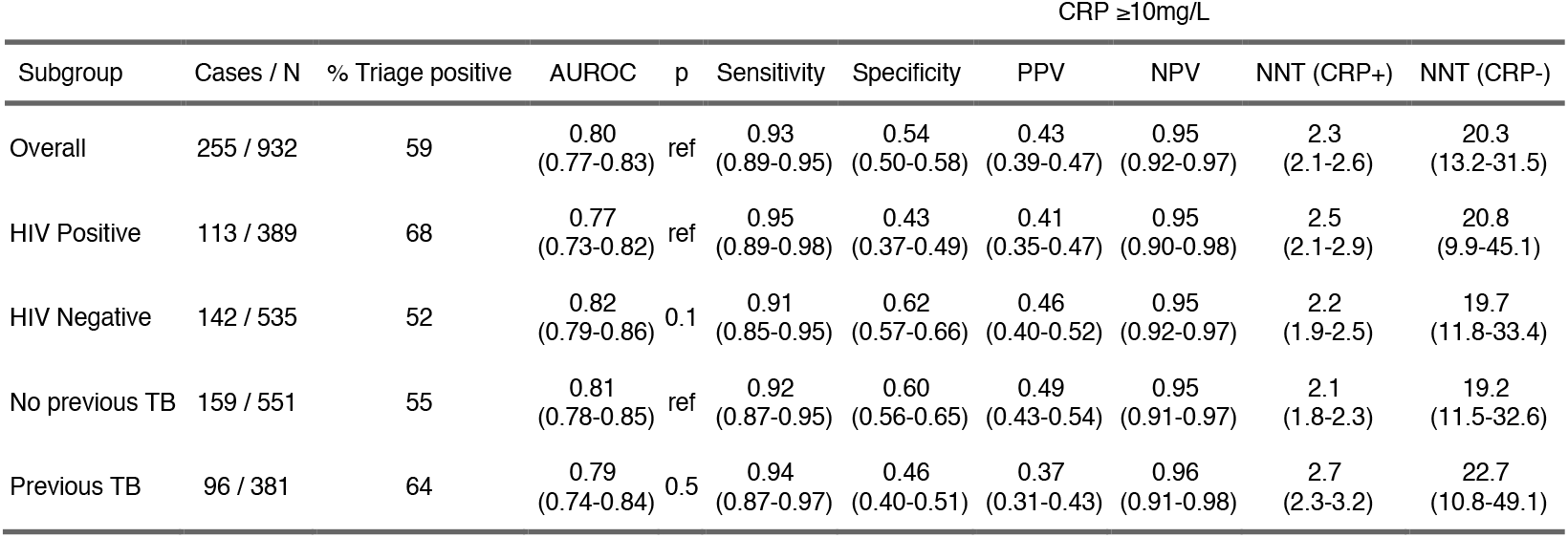
Summary of discriminatory ability of CRP for culture-confirmed pulmonary TB, overall and stratified by key sub-groups of interest. Shown as number TB cases / number individuals (N), % of study population with CRP ≥10mg/L, area under the receiver operator characteristic curve (AUROC), and a p-value from DeLong test for differences across subgroups p. At a threshold of ≥10mg/L, sensitivity, specificity, positive predictive value (PPV) and negative predictive value (NPV) of CRP are shown with 95% confidence intervals, alongside the number needed to test (NNT) with culture to detect one TB case, among participants triage positive (CRP+) and negative (CRP-) at this threshold.

CRP discriminated culture status among people with positive, non-trace Ultra (AUROC 0.75 [95%CI 0.59–0.91]) and had some discriminatory ability for culture status among participants with positive, trace-detected Ultra (AUROC 0.67 [0.48–0.85]), suggesting CRP may help clarify Ultra false-positive results (**Supplementary Figure 3**). The AUROC of CRP for discriminating culture-positive TB among Ultra-negative participants was 0.66 (95%CI 0.57– 0.75; **Figure 4b**). Among the subset of participants with a negative AFB smear the AUROC of CRP was 0.77 (95%CI 0.73–0.81).

**Figure 3:**
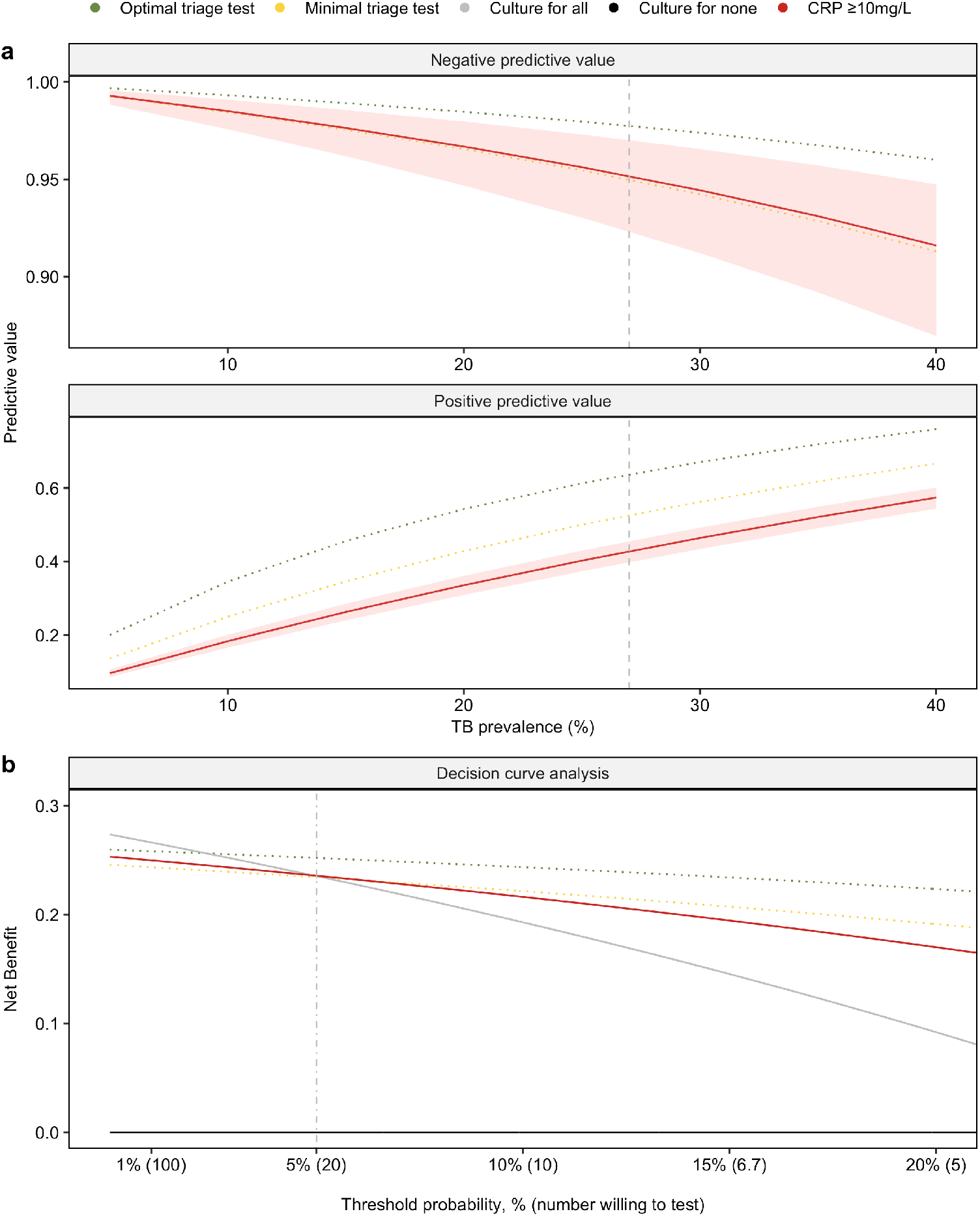
Evaluation of potential clinical utility of CRP as a triage strategy among people presenting with TB-related symptoms. Negative and positive predictive values (a) for CRP for culture-confirmed pulmonary TB using a threshold of ≥10mg/L, a hypothetical ‘optimal’ (sensitivity 95%, specificity 80%), and ‘minimal’ (sensitivity 90%, specificity 70%) tuberculosis triage test as defined by the WHO high-priority target product profiles. Reported across a range of TB prevalences (true prevalence in study population indicated by dashed line [27%]) Decision curve analysis (b) comparing the strategies of an ‘optimal’ or ‘minimal’ triage test, culture for all individuals with CRP ≥10mg/L to a strategy of culture for all or none of the participants meeting study inclusion criteria (i.e. presenting with TB-related symptoms, as defined in text). Dot-dash vertical line represents the threshold probability above which CRP confers net benefit over a ‘culture for all’ strategy.

**Figure 4:**
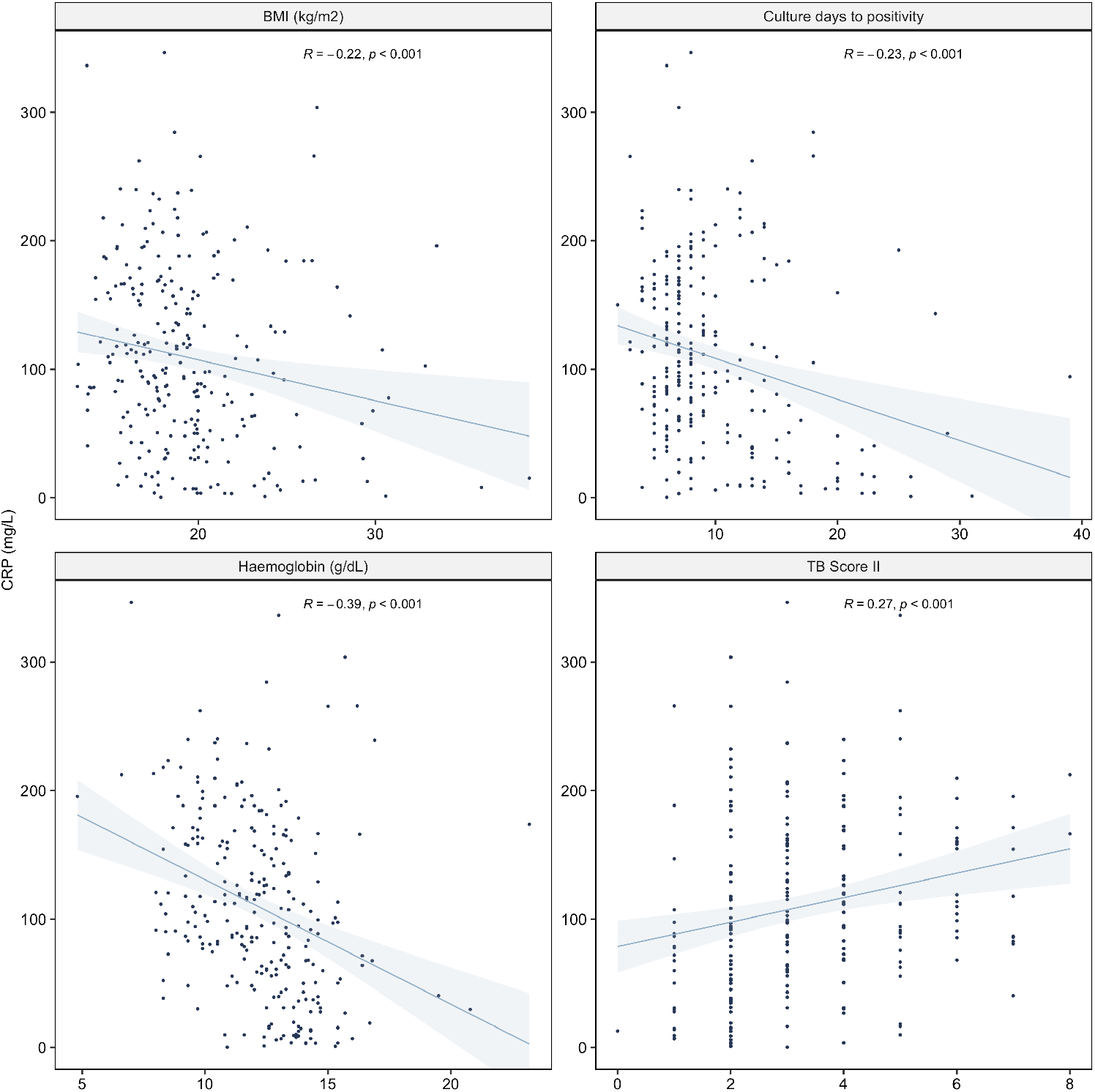
Correlation of CRP with markers of TB severity among culture-confirmed pulmonary TB cases (body mass index ([BMI] culture days to positivity, haemoglobin and TB Score II). TB Score II was only calculated for individuals with complete data for all score components. Spearman rank correlation was calculated.

Among people with culture-confirmed TB, there was strong evidence for an association between higher CRP and indices of higher disease severity (lower BMI, shorter time to culture positivity, lower haemoglobin, and higher TB Score II; all p<0.001; **Figure 4**.

### Clinical utility

To explore the potential clinical utility of CRP as a triage strategy for TB, we first considered the positive and negative predictive values of CRP ≥10mg/L across a range of TB prevalences, benchmarked against a hypothetical optimal and minimal WHO triage test. Among adults attending primary care with TB-related symptoms and prevalences of up to 27%, CRP <10mg/L provided >95% probability that a screened individual does not have TB (95%CI 92–97% at observed TB prevalence; **Table 2** and **Figure 3a**).

We then used decision curve analysis to further assess the clinical utility of CRP-based triage. At the observed TB prevalence (27%), CRP-based triage demonstrated higher net benefit than a strategy of confirmatory testing for all when the number willing to test per true positive TB case detected was up to 20, reflecting a threshold probability of >5% (**Figure 3b**). Above this number willing to test threshold, a strategy of confirmatory testing for all performed best. Net benefit for CRP was similar to the simulated WHO minimal triage test whilst the hypothetical optimal triage test had higher net benefit than a confirmatory test for all strategy when the number willing to test range was up to 33.

### Sensitivity analyses

Sensitivity analyses using a composite microbiological reference standard (Xpert Ultra [including trace] or culture positive TB) resulted in reclassification of 37 primary outcome non-TB cases as having TB. With the composite reference standard or Ultra, diagnostic accuracy of CRP was similar to the primary analysis (**Supplementary Table 5**). Results of decision curve analysis using Ultra as the reference standard were also similar (**Supplementary Figure 5**).

Data on whether participants were able to spontaneously expectorate sputum were only systematically collected for a subset (those recruited after September 2019, n=127). Here, the discriminatory ability of CRP appeared similar among individuals who could not spontaneously produce a sputum sample, compared to those who could, suggesting potential utility for CRP to guide use of sputum induction (**Supplementary Figure 3**).

Illustrative number willing to test ranges where CRP, the hypothetical optimal, or minimal biomarker had higher net benefit than both confirmatory test for all and confirmatory test for none strategies across a range of TB prevalences are presented in **Supplementary Table 5**. The number willing to test range where CRP appeared to confer higher net benefit than confirmatory testing for all or none was wider at lower TB prevalence (estimated range 10– 125 at 5% prevalence compared to 2–14 at 35% prevalence).

## Discussion

In this cohort of 932 adults presenting to primary care with symptoms suggestive of TB, CRP (at a threshold of ≥10mg/L) achieved the minimum sensitivity criteria outlined in the WHO high-priority TPP for a TB triage test, but did not achieve the minimum specificity. The accuracy of CRP was unaffected by HIV or previous TB status, and CRP-based triage demonstrated potential clinical utility to guide confirmatory testing, dependent upon the number of confirmatory tests the health service can perform per TB case detected.

An effective triage test may confer the dual benefits of reducing unnecessary confirmatory testing for people at lower risk of disease, while focusing investigations on those who need them most. However, triage risks missing true positive cases due to imperfect sensitivity.

Using our primary threshold, CRP-based triage would reduce the number of confirmatory tests performed by 41% but would miss 7% of TB cases. Decision curve analysis enabled us to quantify the trade-off between correctly detecting true positive and incorrectly identifying false positive cases. Our findings suggest that the programmatic value of any triage strategy is highly dependent on the number of confirmatory tests the health service is willing to perform per case diagnosed. This number is likely to be context-specific and requires consideration of the resource implications of triage and confirmatory testing, as well as potential costs of missing TB cases. National and international stakeholder engagement are required to further delineate acceptable number willing to test ranges, in order to support further development and implementation of TB triage strategies.

In our setting, CRP-guided triage prior to confirmatory testing offered higher net benefit than a strategy of confirmatory testing for all if the health service is willing to test up to 20 people with a confirmatory test per true positive TB case detected. If the health service can perform more confirmatory tests than this, a confirmatory test for all strategy (i.e., culture or Ultra for all individuals presenting to clinic and meeting WHO symptom criteria for TB evaluation) is likely to perform better. Benchmarking this against the hypothetical optimal and minimal triage test TPPs suggested that the clinical utility of CRP is similar to that of a minimal-standard triage test, whilst an optimal test confers additional net benefit across a wider number willing to test range. Our analyses also suggested that the clinical utility of CRP-guided triage is dependent on TB prevalence and is likely to be useful across a wider number willing to test range in settings with lower TB prevalence, while a strategy of confirmatory testing for all is likely to be preferable when the TB prevalence is very high.

While the discriminatory ability of CRP appeared unaffected by age, HIV status (and ART status among PLHIV), smoking, history of previous TB, age, and BMI, AUROCs were lower among people with anaemia and men. Male sex and anaemia are both associated with later presentation and more severe TB.^21,22^ Our findings of lower discrimination of CRP for TB among these subgroups may reflect lower specificity among more unwell participants, among whom the distribution of CRP is likely to be higher. Notably, CRP was associated with multiple indices of disease severity among TB cases, suggesting that CRP-based triage is more likely to detect more severe cases who may be more likely to transmit TB and experience poor outcomes. CRP offered some discrimination for culture status among participants with positive Ultra (including Ultra trace), suggesting a potential role in resolving false positive results. CRP also demonstrated some discriminatory ability for culture status among Ultra and smear-negative individuals, and, in a subgroup analysis, those who could not spontaneously expectorate sputum, suggesting it may be useful to direct further evaluation among people with negative first-line test results.

Our study is the first, to our knowledge, to report the diagnostic accuracy of CRP for TB in a large, consecutive cohort of outpatients presenting with symptoms compatible with TB with mixed HIV status; and is the first to benchmark CRP-based triage against an Ultra-reference standard. We report higher AUROCs among the HIV-negative subgroup than a recent multi-centre case-control study, providing strong evidence of utility irrespective of HIV status. In the previous study, post-hoc analyses suggested lower accuracy in south-east Asian settings (where participants were recruited at referral centres) compared to populations in southern Africa.^10^ Future studies could further assess the diagnostic accuracy of CRP across a range of settings, including those with lower TB and/or HIV prevalence, and also consider implementation in lower-level health facilities settings where confirmatory testing requires onward referral. We used a robust culture-based reference standard for pulmonary TB (albeit with one, rather than the optimal of two, sputum cultures); with sputum induction for participants unable to produce adequate samples spontaneously. Future studies could also evaluate performance of CRP for extra-pulmonary TB.

There are several limitations to this work. First, CRP was measured retrospectively on stored samples. While we propose that this is a promising tool for point-of-care use, prospective validation using such a platform is required. Previous studies have, however, demonstrated stability of serum CRP over long-term storage and validated quantitative point-of-care CRP against laboratory-based assays.^23,24^ Second, we are unable to characterize the non-TB diagnoses. This, however, reflects the ‘real world’ situation where triage would direct further confirmatory testing and classify people as likely TB or likely non-TB. Third, among included PLHIV, few had a recent CD4 count available, reflecting increasingly infrequent use of routine CD4 counts in the era of ‘test and treat’. This limited our ability to explore the discriminatory ability of CRP across degrees of immunosuppression; previous data suggest sensitivity of CRP for TB may be higher, and specificity lower, among people with lower CD4 counts.^17^ Fourth, it is increasingly recognised that TB symptoms may be minimal: diagnostic algorithms that rely on individuals reporting specific symptoms as an initial gateway may therefore miss cases. Here, we applied a relatively broad definition however future development of triage tests should consider the impact of any ‘pre-screening’ of participants on triage test performance. Finally, we used a complete case analysis, excluding participants with missing CRP results or TB culture results, however, there was no evidence of systematic differences between included and excluded individuals, suggesting low risk of selection bias.

### Conclusion

CRP approached, but did not meet, WHO diagnostic accuracy targets for a triage test to guide confirmatory testing among symptomatic adults in a high TB and HIV-burden setting. Nevertheless, CRP has many desirable operational characteristics and demonstrated potential clinical utility. Additional discriminatory value may be provided through inclusion of CRP in a clinical prediction model, together with other demographic or clinical predictors of risk, or use of CRP in combination with other biomarkers.^25^ Future interventional and health economic studies are required to further evaluate the potential programmatic role of CRP-based triage.

## Data Availability

The de-identified patient data, study protocol, informed consent form, and datasets generated and/or analysed are available from BWPR and CJC on reasonable request.

## Contributors

GT and MN conceived and designed the study. BWPR, TM, ZP, GN and HM were responsible for data collection. CJC and RKG analysed the data with input from BWPR, GT, and MN. CJC produced the figures. CJC and RKG drafted the manuscript. All authors contributed to data interpretation and critically reviewed the manuscript.

## Acknowledgements

We thank the participants, staff of Wallacedene and Scottsdene clinics, and Sisters Charlotte Lawn and Jane Fortuin, and Ms. Danny Sylvester and Ms Zintle Ntwana.

