## Supplementary data for "C-reactive protein as a triage tool for adults with presumptive pulmonary tuberculosis in South Africa: a prospective cohort study"

Claire J Calderwood* (1)

Byron WP Reeve* (2)

Tiffeney Mann (3)

Zaida Palmer (2)

Georgina Nyawo (2)

Hridesh Mishra (2)

Ibrahim Abubakar (1)

Mahdad Noursadeghi (3)

Grant Theron† (2)

Rishi K Gupta† (1)

* Contributed equally

† Contributed equally

**Affiliations**

1. Institute for Global Health, University College London, London, UK

2. DSI-NRF Centre of Excellence for Biomedical Tuberculosis Research; South African Medical Research Council Centre for Tuberculosis Research; Division of Molecular Biology and Human Genetics, Faculty of Medicine and Health Sciences, Stellenbosch University, Cape Town

3. Division of Infection and Immunity, University College London, London, UK

**Corresponding authors**

Dr Rishi K Gupta, PhD

Institute for Global Health, University College London, London, UK

Prof Grant Theron, PhD

DSI-NRF Centre of Excellence for Biomedical Tuberculosis Research; South African Medical Research Council Centre for Tuberculosis Research; and Division of Molecular Biology and Human Genetics, Faculty of Medicine and Health Sciences, Stellenbosch University, Cape Town, South Africa

**Supplementary Figure 1: Sample size required in this study to achieve 90% sensitivity and 70% specificity with a 5% margin of error.**

We used published models for estimates of sample size calculations in diagnostic tests (20) and an anticipated TB prevalence of 30%, based on previous data from the same setting (11). We calculated that a sample size of 326 participants was required to establish whether CRP achieved 90% sensitivity and 70% specificity for TB with ±5% lower bound margin of error. Dashed line represents the sample size achieved this study (n=932).

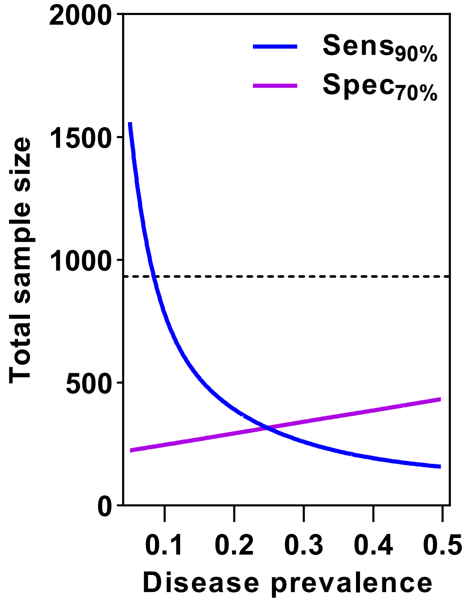

**Supplementary Table 1: Characteristics of participants excluded from analysis.**

Data are n (%) or median (interquartile range). Anaemia was defined as per the World Health Organization: haemoglobin ≤12g/dL for women and <13g/dL for men. Abbreviations: ART: antiretroviral therapy, BMI: body mass index.

| Characteristic | Total | Excluded | Included |
|---|---|---|---|
| **Overall (n = 1097)** | **1,097 (100%)** | **165 (100%)** | **932 (100%)** |
| Scottsdene clinic | 570 (52%) | 73 (44%) | 497 (53%) |
| Wallacedene clinic | 527 (48%) | 92 (56%) | 435 (47%) |
| Culture-positive TB (n = 1070) | 293 (27%) | 26 (19%) | 267 (29%) |
| Age (n = 1072) | 36 (28-47) | 36 (27-46) | 36 (28-47) |
| Female sex (n = 1072) | 502 (47%) | 72 (51%) | 430 (46%) |
| Race (n = 1072) | | | |
| Black | 446 (42%) | 73 (52%) | 373 (40%) |
| Mixed ancestry | 626 (58%) | 67 (48%) | 559 (60%) |
| HIV positive (n = 1063) | 441 (41%) | 52 (37%) | 389 (42%) |
| History of previous TB (n = 1072) | 442 (41%) | 61 (44%) | 381 (41%) |
| BMI (kg/m2) (n = 1064) | 20.3 (18.1-23.9) | 20.3 (18.4-23.8) | 20.3 (18.0-23.9) |
| Anaemia (n = 1053) | 314 (30%) | 39 (30%) | 275 (30%) |
| Presence of cough (n = 1069) | 1,023 (96%) | 129 (93%) | 894 (96%) |
| Presence of fever (n = 930) | 283 (30%) | 35 (30%) | 248 (31%) |
| Presence of weight loss (n = 935) | 554 (59%) | 68 (58%) | 486 (59%) |
| Presence of night sweats (n = 1070) | 760 (71%) | 96 (69%) | 664 (71%) |

**Supplementary Table 2: Summary of diagnostic accuracy of CRP for culture-confirmed pulmonary TB in the overall study population and stratified by key sub-groups of interest, at the maximum Youden index, pre-defined CRP thresholds of ≥10mg/L and >5mg/L, and the threshold which achieved 95% sensitivity.**

Shown as number TB cases / number individuals (N), area under the receiver operator characteristic curve (AUROC), threshold CRP at the maximum Youden index and sensitivity, specificity positive predictive value (PPV) and negative predictive value (NPV) of CRP at thresholds shown.

| | | | | **Max. Youden index** | | | | **CRP ≥10mg/L** | | | | **CRP >5mg/L** | | | | **Sensitivity 95%** | | | |
|---|---|---|---|---|---|---|---|---|---|---|---|---|---|---|---|---|---|---|---|
| Subgroup | Cases / N | AUROC | Threshold CRP | Sensitivity | Specificity | PPV | NPV | Sensitivity | Specificity | PPV | NPV | Sensitivity | Specificity | PPV | NPV | Sensitivity | Specificity | PPV | NPV |
| **Overall** | **255 / 932** | **0.80 (0.77-0.83)** | **27.6** | **0.87 (0.83-0.91)** | **0.66 (0.62-0.69)** | **0.49 (0.45-0.54)** | **0.93 (0.91-0.95)** | **0.93 (0.89-0.95)** | **0.54 (0.50-0.58)** | **0.43 (0.39-0.47)** | **0.95 (0.92-0.97)** | **0.97 (0.94-0.98)** | **0.39 (0.35-0.42)** | **0.37 (0.34-0.41)** | **0.97 (0.94-0.98)** | **0.95 (0.92-0.97)** | **0.50 (0.46-0.53)** | **0.41 (0.38-0.46)** | **0.96 (0.94-0.98)** |
| HIV Positive | 113 / 389 | 0.77 (0.73-0.82) | 63.2 | 0.81 (0.72-0.87) | 0.69 (0.63-0.74) | 0.51 (0.44-0.59) | 0.90 (0.85-0.93) | 0.95 (0.89-0.98) | 0.43 (0.37-0.49) | 0.41 (0.35-0.47) | 0.95 (0.90-0.98) | 1.00 (0.97-1.00) | 0.27 (0.22-0.33) | 0.36 (0.31-0.41) | 1.00 (0.95-1.00) | 0.95 (0.89-0.98) | 0.43 (0.37-0.49) | 0.41 (0.35-0.47) | 0.95 (0.90-0.98) |
| HIV Negative | 142 / 535 | 0.82 (0.79-0.86) | 13.8 | 0.90 (0.84-0.94) | 0.67 (0.62-0.71) | 0.49 (0.43-0.55) | 0.95 (0.92-0.97) | 0.91 (0.85-0.95) | 0.62 (0.57-0.66) | 0.46 (0.40-0.52) | 0.95 (0.92-0.97) | 0.94 (0.89-0.97) | 0.46 (0.41-0.51) | 0.39 (0.34-0.44) | 0.96 (0.92-0.98) | 0.95 (0.90-0.98) | 0.40 (0.36-0.45) | 0.36 (0.32-0.42) | 0.96 (0.91-0.98) |
| No previous TB | 159 / 551 | 0.81 (0.78-0.85) | 29.5 | 0.87 (0.81-0.91) | 0.72 (0.67-0.76) | 0.55 (0.49-0.61) | 0.93 (0.90-0.95) | 0.92 (0.87-0.95) | 0.60 (0.56-0.65) | 0.49 (0.43-0.54) | 0.95 (0.91-0.97) | 0.96 (0.92-0.98) | 0.45 (0.40-0.50) | 0.41 (0.36-0.46) | 0.97 (0.93-0.98) | 0.95 (0.90-0.97) | 0.53 (0.48-0.58) | 0.45 (0.40-0.51) | 0.96 (0.93-0.98) |
| Previous TB | 96 / 381 | 0.79 (0.74-0.84) | 67.4 | 0.73 (0.63-0.81) | 0.76 (0.70-0.80) | 0.50 (0.42-0.59) | 0.89 (0.85-0.93) | 0.94 (0.87-0.97) | 0.46 (0.40-0.51) | 0.37 (0.31-0.43) | 0.96 (0.91-0.98) | 0.98 (0.93-0.99) | 0.30 (0.25-0.36) | 0.32 (0.27-0.38) | 0.98 (0.92-0.99) | 0.95 (0.89-0.98) | 0.45 (0.39-0.51) | 0.37 (0.31-0.43) | 0.96 (0.92-0.98) |

**Supplementary Table 3: Summary of positive and negative predictive value of CRP for culture-confirmed pulmonary TB across a range of TB prevalences.**

Shown as number TB cases / number individuals (N), sensitivity and specificity, and positive predictive value (PPV) and negative predictive value (NPV), with associated confidence intervals.

| | | | | TB prevalence | | | | | | | |
|---|---|---|---|---|---|---|---|---|---|---|---|
| | | | | 10% | | 20% | | 30% | | 40% | |
| Threshold | Cases / N | Sensitivity | Specificity | PPV | NPV | PPV | NPV | PPV | NPV | PPV | NPV |
| Max. Youden index | 255 / 932 | 0.87 (0.83-0.91) | 0.66 (0.62-0.69) | 0.22 (0.20-0.25) | 0.98 (0.97-0.99) | 0.39 (0.35-0.43) | 0.95 (0.94-0.97) | 0.52 (0.48-0.56) | 0.92 (0.89-0.95) | 0.63 (0.59-0.66) | 0.89 (0.84-0.92) |
| Sensitivity 95% | 255 / 932 | 0.95 (0.92-0.97) | 0.50 (0.46-0.53) | 0.17 (0.16-0.19) | 0.99 (0.98-0.99) | 0.32 (0.30-0.34) | 0.98 (0.96-0.99) | 0.45 (0.42-0.47) | 0.96 (0.93-0.98) | 0.56 (0.53-0.58) | 0.94 (0.89-0.96) |
| CRP ≥10mg/L | 255 / 932 | 0.93 (0.89-0.95) | 0.54 (0.50-0.58) | 0.18 (0.17-0.20) | 0.98 (0.98-0.99) | 0.34 (0.31-0.36) | 0.97 (0.95-0.98) | 0.46 (0.43-0.49) | 0.94 (0.91-0.97) | 0.57 (0.54-0.60) | 0.92 (0.87-0.95) |
| CRP >5mg/L | 255 / 932 | 0.97 (0.94-0.98) | 0.39 (0.35-0.42) | 0.15 (0.14-0.16) | 0.99 (0.98-1.00) | 0.28 (0.27-0.30) | 0.98 (0.96-0.99) | 0.40 (0.38-0.42) | 0.97 (0.93-0.98) | 0.51 (0.49-0.53) | 0.95 (0.90-0.98) |

**Supplementary Figure 2: Diagnostic accuracy of CRP for culture-confirmed pulmonary TB, stratified by additional subgroups of interest**

Area under the receiver operator characteristic curve (AUROC) of CRP to distinguish TB versus non-TB diagnoses, (a) in whole study cohort and (b) among people living with HIV. Anaemia was defined as per WHO: haemoglobin ≤12g/dL for women and <13g/dL for men. BMI was classified using WHO definitions: underweight BMI <18, normal BMI 18-24.9 and overweight BMI ≥25. TB Score II severity categories defined as previously (13) and was restricted to study population with data available for all components. On DeLong tests, AUROC appeared inferior among male, as compared to female participants ($p = 0.03$), and among people with anaemia compared to those without ($p = 0.003$). There was no evidence for differences across the other subgroups shown ($p \geq 0.1$ across all).

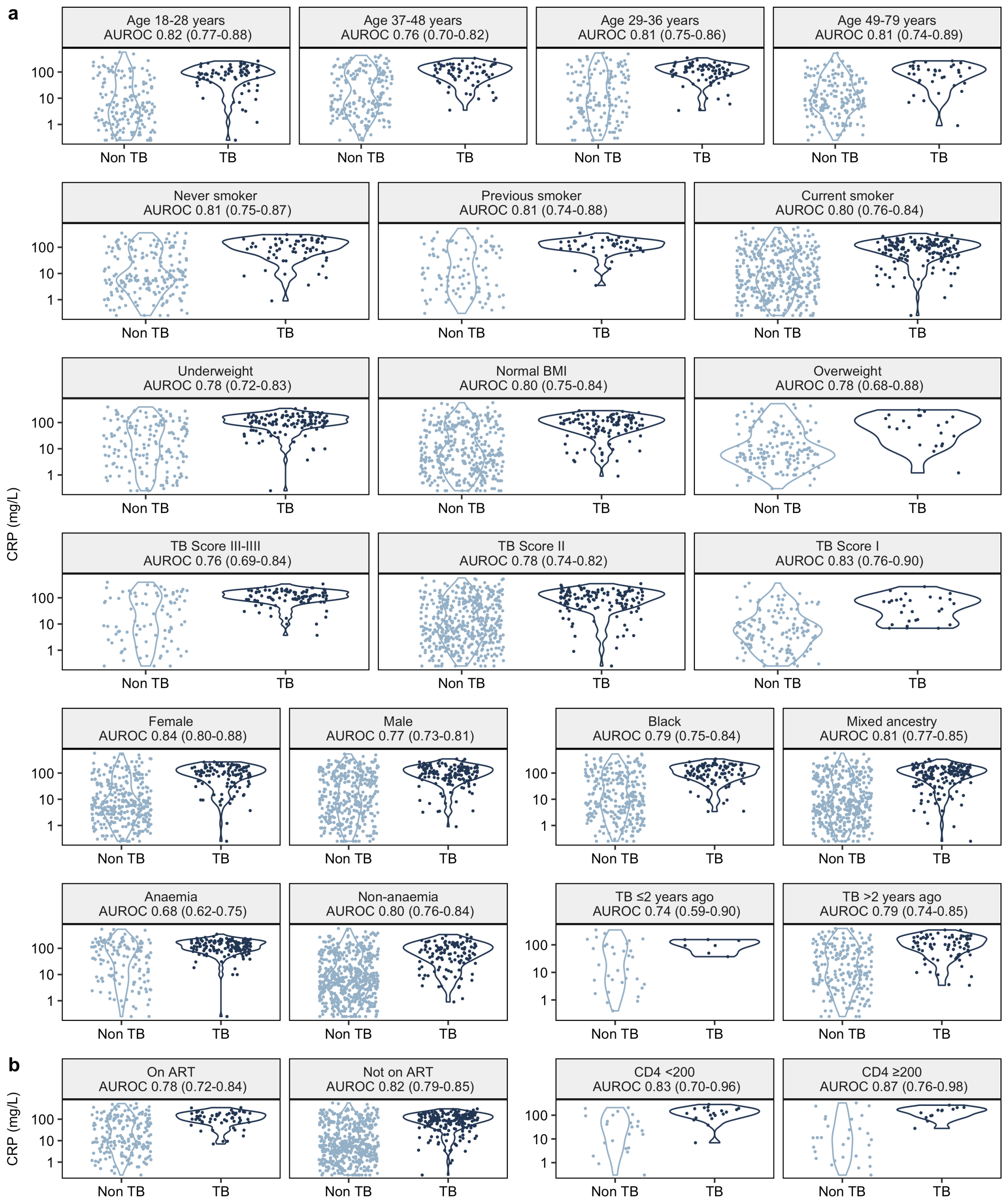
a
Age 18-28 years AUROC 0.82 (0.77-0.88)
Age 37-48 years AUROC 0.76 (0.70-0.82)
Age 29-36 years AUROC 0.81 (0.75-0.86)
Age 49-79 years AUROC 0.81 (0.74-0.89)
Never smoker AUROC 0.81 (0.75-0.87)
Previous smoker AUROC 0.81 (0.74-0.88)
Current smoker AUROC 0.80 (0.76-0.84)
Underweight AUROC 0.78 (0.72-0.83)
Normal BMI AUROC 0.80 (0.75-0.84)
Overweight AUROC 0.78 (0.68-0.88)
TB Score III-IIII AUROC 0.76 (0.69-0.84)
TB Score II AUROC 0.78 (0.74-0.82)
TB Score I AUROC 0.83 (0.76-0.90)
Female AUROC 0.84 (0.80-0.88)
Male AUROC 0.77 (0.73-0.81)
Black AUROC 0.79 (0.75-0.84)
Mixed ancestry AUROC 0.81 (0.77-0.85)
Anaemia AUROC 0.68 (0.62-0.75)
Non-anaemia AUROC 0.80 (0.76-0.84)
TB ≤2 years ago AUROC 0.74 (0.59-0.90)
TB >2 years ago AUROC 0.79 (0.74-0.85)
b
On ART AUROC 0.78 (0.72-0.84)
Not on ART AUROC 0.82 (0.79-0.85)
CD4 <200 AUROC 0.83 (0.70-0.96)
CD4 ≥200 AUROC 0.87 (0.76-0.98)
CRP (mg/L)
100
10
1
Non TB
TB

**Supplementary Table 5: Post-hoc analysis of sensitivity and specificity of CRP at a threshold of ≥10mg/L for culture-confirmed pulmonary TB across strata of sex and anaemia status**

Shown as number TB cases / number individuals (N), area under the receiver operator characteristic curve (AUROC) and sensitivity, specificity, positive predictive value (PPV) and negative predictive value (NPV) at a CRP threshold of ≥10mg/L, with 95% confidence intervals.

| | | | | CRP ≥10mg/L | | |
|---|---|---|---|---|---|---|
| Subgroup | Cases / N | AUROC | Sensitivity | Specificity | PPV | NPV |
| Female | 110 / 430 | 0.84 (0.80-0.88) | 0.91 (0.84-0.95) | 0.63 (0.58-0.68) | 0.46 (0.39-0.53) | 0.95 (0.92-0.97) |
| Male | 145 / 502 | 0.77 (0.73-0.81) | 0.94 (0.89-0.97) | 0.46 (0.41-0.51) | 0.41 (0.36-0.47) | 0.95 (0.90-0.97) |
| Anaemia | 136 / 275 | 0.68 (0.62-0.75) | 0.98 (0.94-0.99) | 0.29 (0.23-0.38) | 0.58 (0.51-0.64) | 0.93 (0.82-0.98) |
| Non-anaemia | 118 / 650 | 0.80 (0.76-0.84) | 0.86 (0.79-0.91) | 0.61 (0.56-0.65) | 0.33 (0.28-0.38) | 0.95 (0.92-0.97) |

**Supplementary Figure 3: Discriminatory ability of CRP for culture-confirmed pulmonary TB among adults presenting with TB-related symptoms, stratified by Xpert Ultra result, and among smear negative individuals.**

Area under the receiver operator characteristic curve (AUROC) of CRP to distinguish TB versus non-TB diagnoses in (a) Ultra-positive individuals, stratified by trace or higher-grade result, (b) Ultra negative individuals and (c) smear negative individuals. Individual points are coloured by whether there was a history of previous TB (dark blue) or not (light blue).

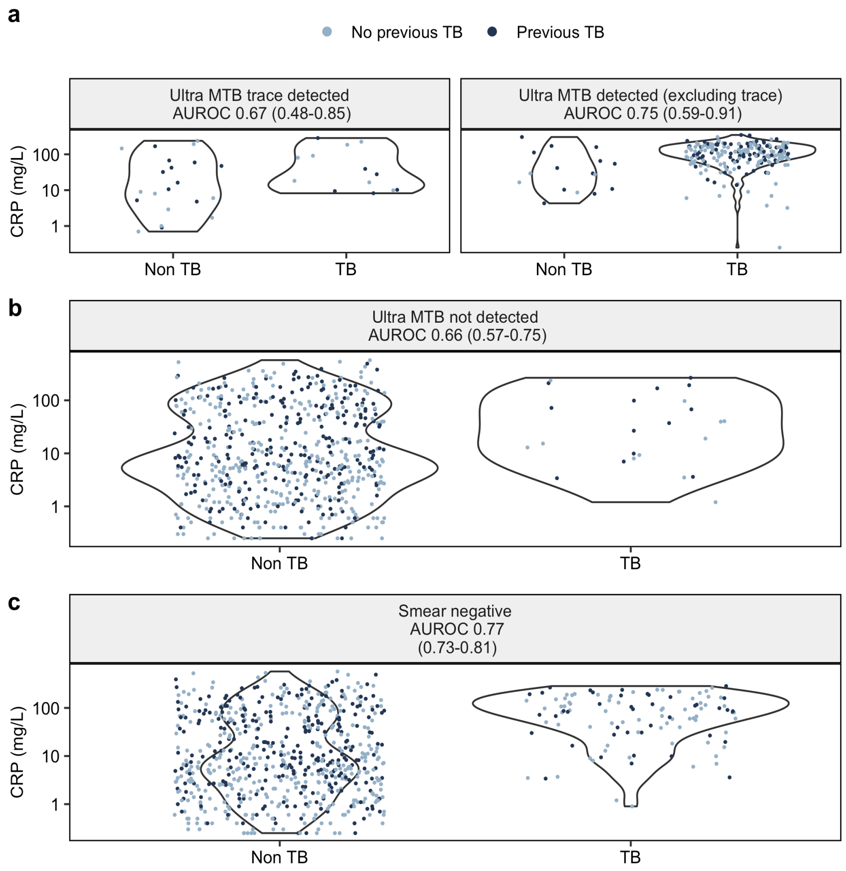

**Supplementary Figure 4: Discriminatory ability of CRP for culture-confirmed pulmonary TB, stratified by whether participants were able to produce a spontaneous sputum sample (data available for n = 127)**

AUROC = Area under the receiver operator characteristic curve.

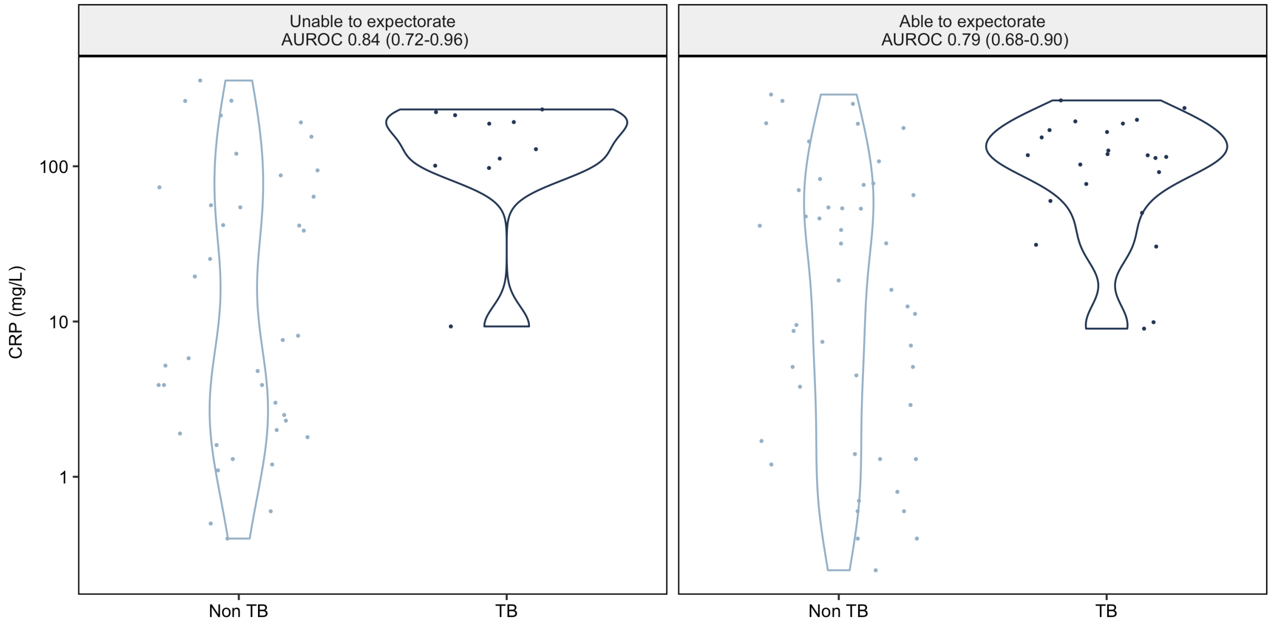

**Supplementary Table 5: Sensitivity analyses - discriminatory ability of CRP across varying reference standard definitions for TB.**

Shown as TB cases / number of individuals with result available for reference standard shown; area under the receiver operator characteristic curve (AUROC) and associated 95% confidence interval (95% CI); and sensitivity and specificity, positive (PPV) and negative (NPV) predictive value at a prespecified ≥10mg/L threshold

| | | | | | CRP ≥10mg/L | | | |
|---|---|---|---|---|---|---|---|---|
| Outcome | Subgroup | Cases / N | % Triage positive | AUROC | Sensitivity | Specificity | PPV | NPV |
| **Primary outcome: Culture+ TB** | **Overall** | **255 / 932** | **59** | **0.80 (0.77-0.83)** | **0.93 (0.89-0.95)** | **0.54 (0.50-0.58)** | **0.43 (0.39-0.47)** | **0.95 (0.92-0.97)** |
| Ultra/Culture+ reference | Overall | 292 / 932 | 59 | 0.78 (0.75-0.81) | 0.89 (0.85-0.92) | 0.55 (0.51-0.59) | 0.48 (0.43-0.52) | 0.92 (0.89-0.94) |
| Ultra+ reference | Overall | 260 / 896 | 61 | 0.79 (0.76-0.82) | 0.91 (0.87-0.94) | 0.55 (0.51-0.59) | 0.45 (0.41-0.49) | 0.94 (0.91-0.96) |

**Supplementary Table 7: Sensitivity and Specificity of Xpert MTB/RIF Ultra observed in this study**

| Category | Cases / N | Sensitivity | Specificity |
|---|---|---|---|
| Xpert MTB/RIF Ultra | 247 / 896 | 0.90 (0.86-0.94) | 0.94 (0.92-0.96) |

**Supplementary Table 8: Sensitivity analyses - Number willing to test (NWT) range at which CRP offers clinical utility over a 'test all' and 'test none' strategy, across varying TB prevalence.**

| | NWT range across TB prevalence | | | |
|---|---|---|---|---|
| Strategy | 5% | 15% | 25% | 35% |
| CRP ≥10mg/L | 10 - 125 | 4 - 42 | 2 - 23 | 2 - 14 |
| Optimal WHO triage test | 5 - 250 | 2 - 83 | 2 - 48 | 1 - 29 |
| Minimal WHO triage test | 7 - 125 | 3 - 38 | 2 - 21 | 2 - 14 |

**Supplementary Figure 5: Decision curve analysis with Ultra MTB/RIF reference standard.**

Comparing the strategies of an 'optimal' or 'minimal' triage test, and Ultra for all individuals with CRP ≥10mg/L to a strategy of Ultra for all or none of the participants meeting study inclusion criteria.

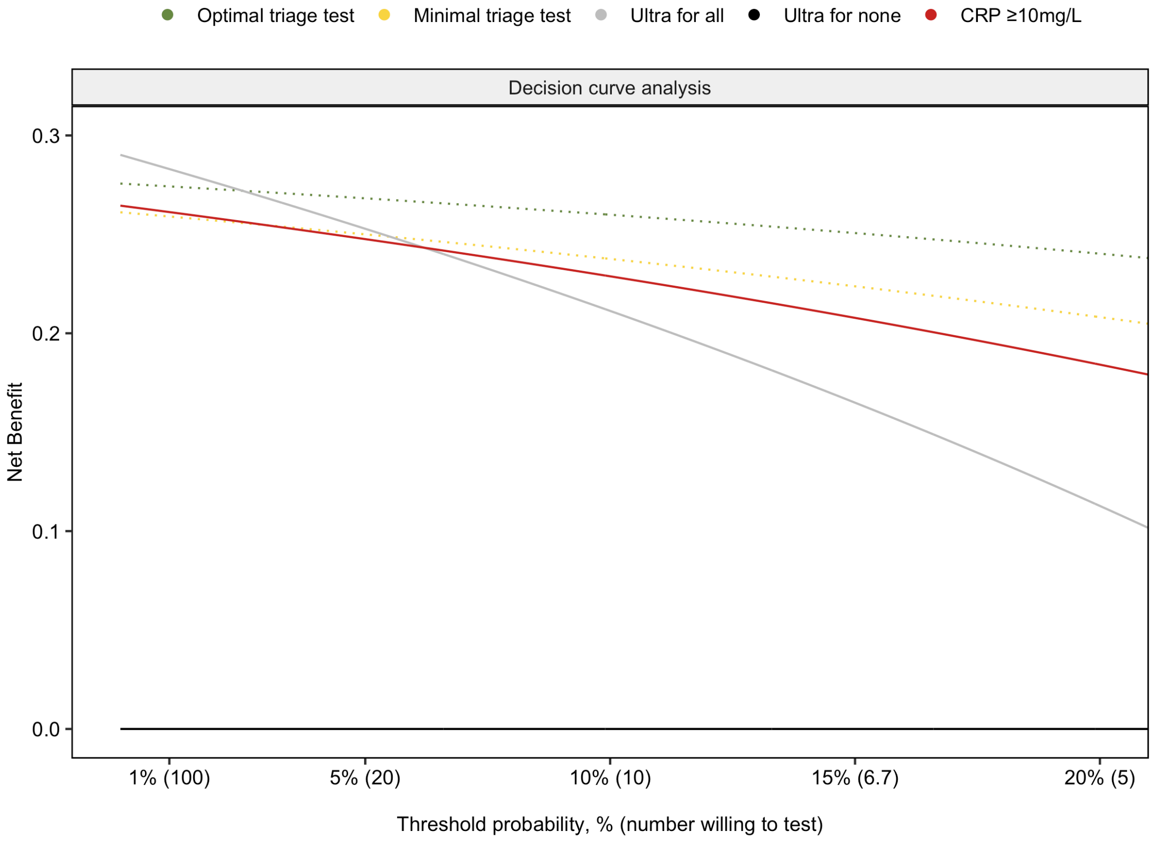
